# Adolescent videogaming, cognitive development and interactions with gender in the SCAMP cohort

**DOI:** 10.64898/2026.09.15.26363113

**Authors:** Rhiannon Thompson, Joel Heller, Nicole Curtis, Sara Rudbæk Larsen, Chen Shen, Rachel B. Smith, Michael S.C Thomas, Iroise Dumontheil, Mireille B. Toledano

## Abstract

**Background:** There is evidence that non-problematic videogaming, particularly action and first-person shooter gaming, is associated with enhanced cognition in some areas such as attention and spatial cognition. However, many studies to date have been cross-sectional, making it difficult to determine how videogaming influences cognitive development during adolescence.

**Objectives:** To investigate whether videogaming habits in early adolescence, including gaming genre, are associated with cognitive development in the prospective Study of Cognition, Adolescents and Mobile Phones (SCAMP).

**Methods:** Data on videogaming habits, demographics and cognitive abilities (executive functioning, visual attention, mental rotation and sustained attention) was collected at two timepoints, when participants were age 11-12 (baseline, N= 6590) and 13-15 (follow-up, N= 4976). Multi-level linear regression, adjusted for demographics and baseline cognition, was used to evaluate whether baseline videogaming (hours per week and specific genres) predicted cognitive abilities cross-sectionally and at follow-up. Interaction terms and stratified analyses were used to examine potential gender differences.

**Results:** Overall videogaming duration at baseline was not associated with subsequent cognitive development, however playing some specific genres was associated with improved mental rotation (first-person shooter and strategy games) and sustained and visual attention (platform games), and some genres (first-person shooter and roleplayer games) were associated with worse sustained attention. Further, effects were modified by gender, with stratified analyses suggesting detrimental effects of overall gaming on sustained attention in boys, and improvements in visual attention in girls.

**Conclusions:** Overall videogaming duration was not consistently associated with cognitive development during early adolescence. However, associations varied according to participant gender and gaming genre, supporting the hypothesis that videogaming should not be considered a homogeneous activity. These findings highlight the importance of distinguishing between specific gaming experiences and broader screen-time exposure when evaluating potential impacts on adolescent cognitive development and informing public health recommendations.

## Introduction

The cognitive impacts of videogaming, a common recreational activity for adolescents worldwide, has been extensively researched but findings remain mixed (1-3). Adolescence is a period of rapid cognitive development where behavioural and environmental factors can exert a large influence on outcomes (4, 5). Individual studies and meta-analyses have suggested that videogaming may be associated with improvements in attention, working memory visuospatial processing, cognitive control and inhibition, task-switching, and processing speed, particularly for action and first-person shooter (FPS) games (1, 6-8). There is corresponding evidence from magnetic resonance imaging studies that videogaming is associated with beneficial effects on grey matter, brain activity, and functional connectivity (9) and signal differences in cognitive task-related regions (6).

Although this evidence suggests positive differences in brain functioning and cognition in adolescents who play videogames, many studies are cross-sectional, involve relatively small samples, or focus on specific groups of gamers (habitual action video-gamers, often healthy, younger males) (1, 7). As the cognitive and brain changes undergone at each developmental stage vary in complex ways, longitudinal population-based research offers the opportunity to determine the impacts of videogaming at different ages on later cognitive development whilst accounting for baseline cognition, enhancing causal inference.

Intervention studies have provided some evidence that action-game training can improve specific cognitive skills, namely top-down attention and spatial cognition, supporting potential causal interpretation (1, 7). However, Hilgard and colleagues argued that the action-gaming literature may overstate cognitive benefits because of publication bias, overlapping samples and salami-slicing of outcomes into different papers. Their reanalysis of a highly cited meta-analysis revealed little to no evidence of cognitive effects once publication bias was accounted for (10). Given current biases in the literature, research that helps untangle these relationships is warranted, particularly longitudinal studies that analyse a range of outcomes and present all results together (including null effects).

Further, there is evidence that videogaming addiction and problematic (e.g. excessive) videogaming may be associated with detrimental impacts on cognition, especially in young people, which could be related to the displacement of health-forming behaviours like sleep, socialising and exercise (11-14). Therefore, in addition to the presence and duration of play, contextual factors like timing (e.g. weekend versus weekday play, social versus isolated play) may also be important indicators, and research should explore non-linear patterns of association. Importantly, videogaming is not a homogenous activity, as there exists substantial variability in content and skills employed across different genres. Although research has explored the impact of whether and how much young people play, more research is required to explore differences in play such as genre and timing.

Further, boys are generally more likely to play video games, and there’s also evidence for gender differences in engagement habits like genre, choice of device, time spent playing, and frequency of play (7, 15). An experimental study by Feng and colleagues found that action gaming reduced gender differences in spatial cognition, supporting the hypothesis that videogaming could have differential impacts on cognition by gender and could particularly benefit girls (8). Yet, few studies explore whether the association between videogaming and adolescent cognitive development is moderated by gender, especially large prospective studies.

The Study of Cognition, Adolescents and Mobile Phones (SCAMP) is a large London-based prospective longitudinal cohort study investigating the impacts of digital technology on adolescent psychological development, with rich data on a range of cognitive domains. The aim of this study was to investigate whether videogaming (duration and genre) in early adolescence (baseline age 11–12) predicted subsequent cognitive development (age 13–15), and whether these effects were moderated by gender. We hypothesised that action and first-person shooter games would be associated with beneficial cognitive outcomes, whereas higher overall gaming exposure may show adverse associations.

## Methods

### Participants

SCAMP is a longitudinal cohort study about digital technology use and psychological development in adolescents, of which a detailed protocol has been published previously (16). Briefly, 6590 participants living in and around Greater London were recruited and took part via their schools in year 7 (aged 11–12 during 2014–2016), completing computer-based batteries containing cognitive tasks and questionnaires using the Psytools (Delosis) software. A subset of 4976 participants then completed a follow-up assessment in years 9-10 (aged 13–15 in 2016–2018). In this study, participants with complete data on selected demographics, videogaming behaviours, and cognitive outcomes were included in the analytical sample.

### Measures

#### Videogaming behaviour

Participants were asked the duration of time spent playing videogames per day (any games, and first-person shooter games specifically) on weekdays and on weekends (Never, 1-10 minutes, 11-30 minutes, 31-59 minutes, 1-2 hours, 3-4 hours, 5+ hours), which was averaged to compute average weekly gaming duration as the main exposure variable. Participants were also asked how often they played a range of game genres (never, occasionally or often), each provided with one or more examples as follows:

- First-person shooter, e.g. Call of Duty, Medal of Honor
- Action, e.g. Tomb Raider, Grand Theft Auto
- Multiplayer online, e.g. Shadowrun Online
- Sport, e.g. FiFA, Pro Wrestling
- Racing e.g. Grid, Trackmania
- Role-playing e.g. Skyrim, Wasteland
- Puzzles e.g. Candy Crush
- Strategy e.g Starcraft, Command and Conquer
- Flight simulator
- Platform e.g Super Mario, Contrast,
- Nintendo wii/Xbox Kinect/PlayStation Move
- Other e.g. educational or learning games

### Cognitive measures

Cognitive measures were taken at baseline and follow-up. An executive function composite z-score (reflecting cognitive flexibility, working memory, and inhibition) was created using time taken to complete the Trail Making Test (TMT, reverse coded)(17), estimated span on the Backwards Digit Span (BDS)(18), and total number of errors on the Spatial Working Memory (SWM, reverse coded) tasks (18). Visual attention was assessed using accuracy on the Enumeration task (19). Sustained attention was assessed by the AX-CPT continuous performance task, using the d-prime measure (correct hits/commission errors)(20). Mental rotation was assessed using the number of correct responses in the Mental Rotation Task(21). Higher scores on all outcomes reflected better performance.

### Sociodemographic covariates

Based on baseline survey responses and school meta-data, participant gender (male, female), age (continuous, based on date of birth), ethnicity (based on specific categories, compound variables for white, black, Asian, other or mixed ethnicity), school type (state/independent) and socioeconomic status (parental National Statistics Socio-economic Classification (NS-SEC), composite three class version, highest of any parent) were included as covariates, selected a priori. The inclusion of self-reported average sleep duration at baseline did not improve model fit.

### Statistical analysis

Chi-squared tests were used to compare gender differences in playing different game genres at baseline. To estimate total weekly gaming duration, a weekly average was computed from weekend and weekday play, taking the category midpoint as the continuous daily value and weighting weekday use at 5/7 and weekend use at 2/7. Multi-level linear regression was conducted evaluating the associations between gaming (weekly gaming duration overall and on first-person shooter games; and frequency of playing specific gaming genres) and cognitive performance (i) cross-sectionally using baseline gaming and cognitive performance, (ii) longitudinally using baseline gaming and follow-up cognitive performance and (iii) longitudinally using the mean weekly videogaming duration between baseline and follow-up (not possible for gaming genre which are categorical data) and follow-up cognitive performance. Complete cases analyses were conducted, where participants were included in a given model if they had the complete set of required videogaming, outcome and covariate data. Models were adjusted for gender, age, ethnicity, socioeconomic status, and school type (state/independent). Longitudinal models were further adjusted for cognition at baseline (performance on the outcome task) and time difference between assessments. Multi-level analyses were run with a random intercept for school to account for clustering, unless a single-level model (without school clustering) produced a better fit in which case simple linear models were run. To take into account the issue of multiple comparisons, False Discovery Rate (FDR) corrected p-values were estimated and presented alongside the individual models’ *p*-values.

Models estimating the association between continuous baseline gaming and cognition were then re-run including an interaction term for gender, and stratified analyses were run to explore any significant effects *(p* < 0.05). Finally, to explore whether specific choice of genre helped explain any gender differences, gender stratified models were adjusted for game genre, including the categorical variable (never, occasionally, often) for any genres that were significantly associated with both gender and the cognitive outcome (models run independently adjusted for each genre).

Sensitivity analyses were run (i) estimating the impact of weekend and weekday gaming as separate exposures in separate models, and (ii) estimating weekly duration of gaming as a categorical exposure to explore non-linearity (Never, 1-30 minutes, 31 minutes-2 hours, 3+ hours).

### Ethical considerations

The SCAMP study and its amendments were approved by the North-West Haydock Research Ethics Committee (14/NW/0347). Head teachers consented for their schools to take part. Students and their parents received written information about the study in advance and were given an opportunity to opt-out.

### Public involvement and engagement

This work is part of the SCAMP Research Challenge programme, launched in October 2022. The SCAMP Research Challenge is an innovative approach to getting young people involved in scientific research. Teams of students at participating schools proposed research projects and helped collect data (not analysed in this study). A team of students from Bishopshalt School (Uxbridge) were involved in idea formulation, the analytical plan, and reviewing literature in workshops with SCAMP researchers as the project was developed.

## Results

A total of 6590 participants (Table 1) completed baseline assessment (mean age 11.62 years, 52% female) and 4976 participants took part in follow-up assessment (mean age 13.81, 54% female). Participants required complete gaming, outcome and covariate data to be included in analyses, so the analytical sample ranged from 366 and 4818 depending on timepoint, gaming and cognitive outcome data (particularly varying completion rates by task, especially for longitudinal analyses where a task needed to be completed at both timepoints). For all game types but platform games (Figure 1), chi-squared tests indicated statistically significant gender differences in how often these games were played (*p* < 0.01).

**Table 1.** Descriptive statistics of participant gaming behaviours, cognition and demographic characteristics.

|  | Baseline (N = 6,590) | Follow-up (N = 4,976) |
| --- | --- | --- |
| **Weekly gaming duration (hours)** |  |  |
| Mean(range) | 1.06 (1,5) | 1.12 (0,5) |
| Missing (%) | 64 (0.97) | 112 (2.3) |
| **Age** |  |  |
| Mean (range) | 11.62 (9.08, 14.1) | 13.81 (12.07, 15.11) |
| Missing (%) | 8 (0.12) | 7 (0.14) |
| **Gender** |  |  |
| Male (%) | 3,128 (47.5) | 2,272 (45.7) |
| Female (%) | 3,458 (52.5) | 2,704 (54.3) |
| Missing (%) | 4 (0.06) | 0 (0) |
| **Ethnicity** |  |  |
| White | 2,710 (41.1) | 725 (14.6) |
| Asian | 1,712 (26.0) | 1,327 (26.7) |
| Black | 979 (14.9) | 748 (15.0) |
| Mixed or Other | 764 (11.6) | 521 (10.5) |
| Missing (%) | 425 (6.5) | 1,655 (33.3) |
| **Highest parental NSSEC** |  |  |
| Managerial and professional occupations | 3,264 (49.5) | 2,644 (53.1) |
| Intermediate occupations | 1,390 (21.1) | 989 (19.9) |
| Routine and manual occupations | 962 (14.6) | 569 (11.4) |
| Missing (%) | 974 (14.8) | 774 (15.6) |
| **School type** |  |  |
| State (%) | 5,160 (78.3) | 3,689 (74.1) |
| Independent (%) | 1,430 (21.7) | 1,287 (25.9) |
| **Executive functioning** |  |  |
| Median (IQR) | 0.04 (-4.18, 2.09) | 0.22 (-2.4, 2.66) |
| Missing (%) | 831 (12.61) | 680 (13.67) |
| **Visual attention (enumeration)** |  |  |
| Median (IQR) | 1.97 (1, 6.95) | 2.01 (1, 6.99) |
| Missing (%) | 2,693 (40.86) | 1675 (33.66) |
| **Mental rotation** |  |  |
| Median (IQR) | 8.37 (2, 15) | 8.79 (2, 16) |
| Missing (%) | 3,462 (52.53) | 3,366 (67.64) |
| **Sustained attention (AX-CPT)** |  |  |
| Median (IQR) | 2.8 (-0.62, 3.94) | 3.03 (-0.61, 3.94) |
| Missing (%) | 5,024 (76.24) | 4085 (82.09) |

**Figure 1.**
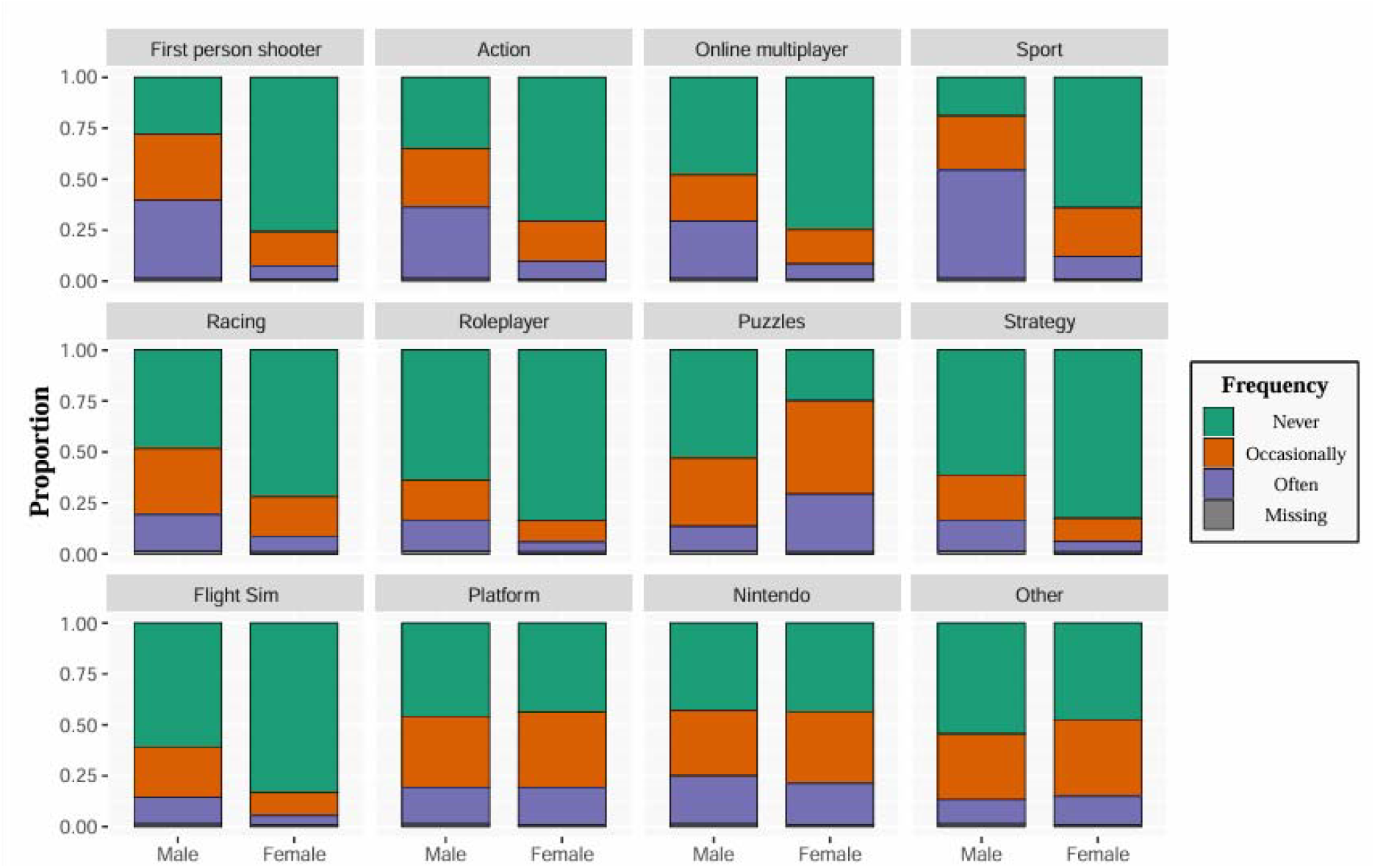
Frequency of playing different gaming genres by gender

At baseline (Figure 2, Appendix A), statistically significant cross-sectional associations were not observed for the weekly duration of play overall with any cognitive measure, however increased weekly duration of playing first-person shooter games, and frequency of playing first-person shooter games, action games, and flight simulator games were associated with worse executive functioning, whereas strategy and puzzle games were associated with better executive functioning; playing racing, Nintendo wii/Xbox Kinect/PlayStation Move and other games occasionally was associated with better executive functioning, but playing them often was associated with worse executive functioning (all associations survived FDR correction). Playing multiplayer games occasionally was associated with better visual attention (surviving FDR correction). Playing sports games was associated with worse mental rotation, as was playing Nintendo wii/Xbox Kinect/PlayStation Move or action games ‘often’. Playing action games often was associated with worse sustained attention, but occasional play of racing and other games was associated with better sustained attention.

**Figure 2.**
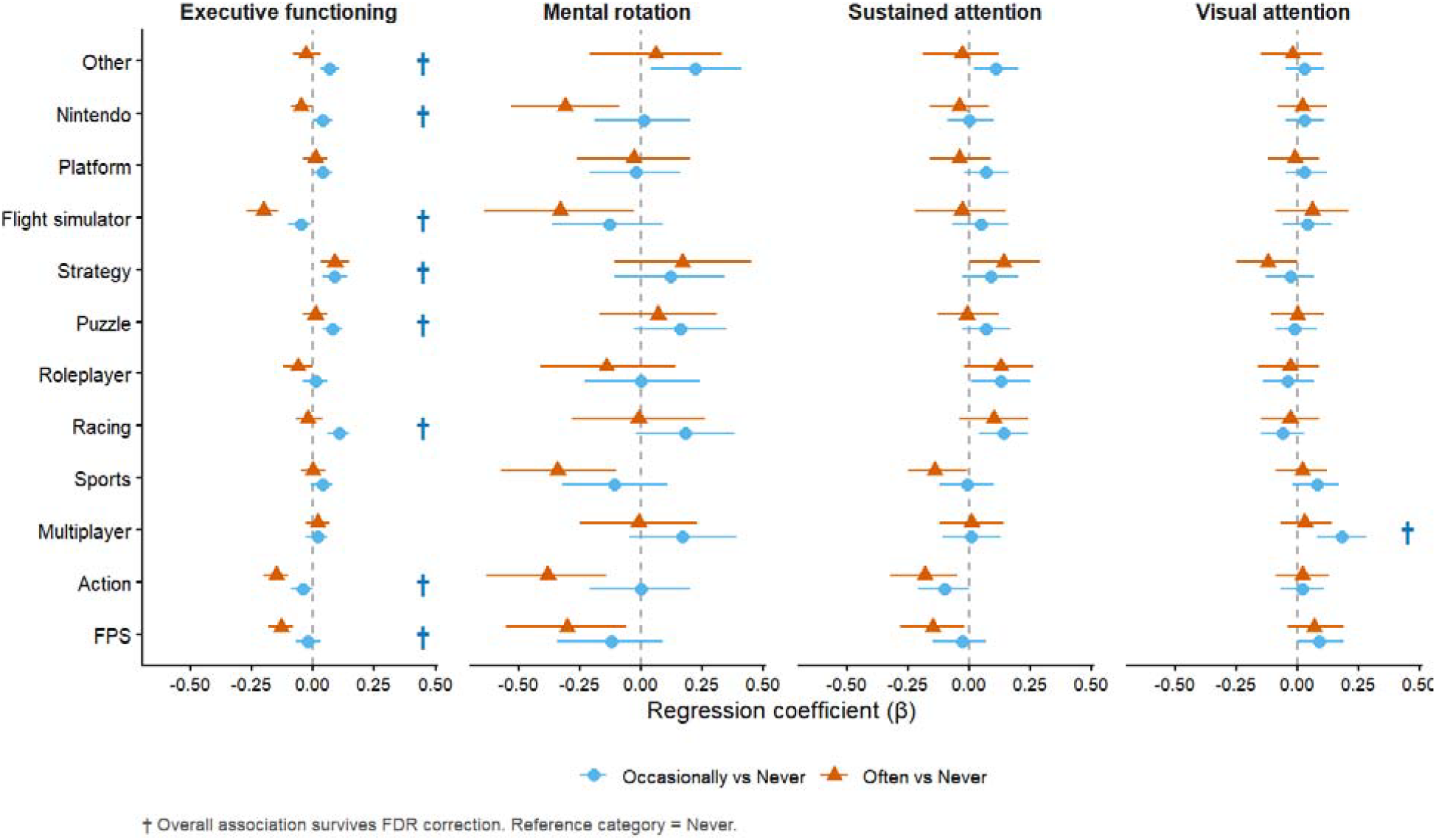
Cross-sectional associations between playing different videogaming genres and cognitive abilities at baseline

Longitudinally (Figure 3, Appendix B), playing platform games occasionally, but not often, was associated with improvements in visual attention and playing first-person shooter games occasionally, but not often, was associated with improvements in mental rotation, as was playing strategy games occasionally or often (with often exerting a stronger effect). Weekly duration of playing first person shooter games was associated with worse sustained attention at follow-up, as was playing ‘often’, as well as ‘often’ playing role-playing games. Often playing platform games was associated with better sustained attention at follow-up. However, none of the above longitudinal associations were significant after FDR correction.

**Figure 3.**
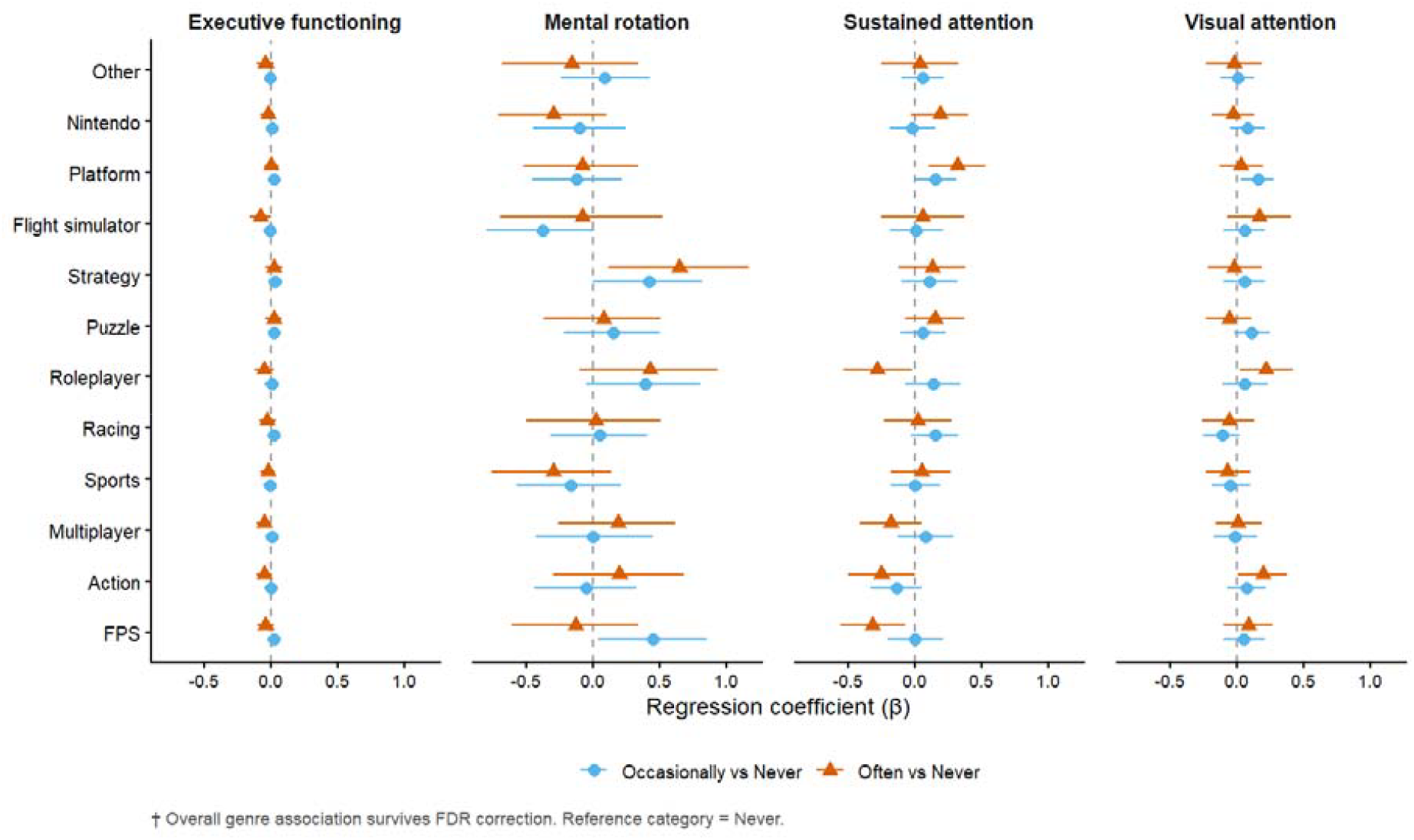
Longitudinal associations between playing different videogaming genres at baseline and cognitive abilities at follow-up

Associations between the weekly duration of play at baseline and follow-up visual attention and sustained attention were significantly moderated by gender (Figure 4, Appendix D). In boys, visual attention and gaming were not associated, whereas weekly duration of play was associated with better visual attention in girls, remaining statistically significant after adjustment for all gaming genres except action and role-playing (although the direction and magnitude of effect was consistent). Weekly duration of play at baseline was associated with worse sustained attention at follow-up in boys, surviving adjustment for some game genres with a consistent direction and magnitude of association across models. Overall, sustained attention at follow-up was not associated with baseline duration of play in girls, but significant associations emerged when adjusting for action, multiplayer, racing and role-playing gaming, with baseline duration of play predicting better follow-up sustained attention. This suggests that the relationship between gaming duration and attention may partly depend on the types of games played, but this does not totally explain differences by gender.

**Figure 4.**
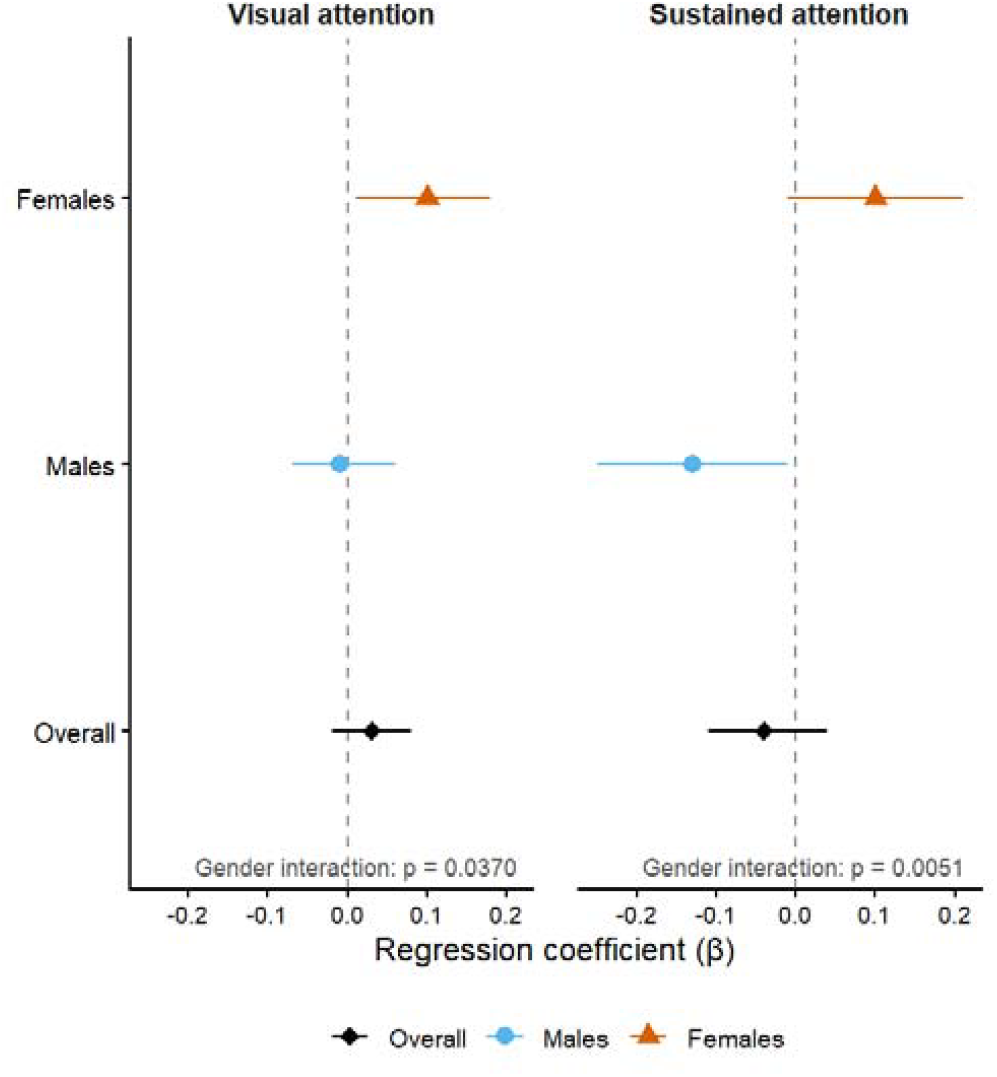
Longitudinal associations between baseline weekly videogaming duration and follow-up cognitive performance by gender

### Sensitivity analyses

Results were broadly equivalent for weekday and weekend gaming duration, with little apparent association between overall videogaming and cognition. However, the association with improvements in visual attention in girls and worsening sustained attention in boys was only significant for weekend play (Appendix E).

Examining videogaming duration categorically (Appendix F), results were consistent with the main findings, with the duration of first-person shooter gaming associated with sustained attention and the strongest association at 3+ hours per week. However, a positive non-significant effect of playing 30–120 minutes total gaming duration per week and executive functioning was observed (β = 0.09, 95% CI: 0.01, 0.17) relative to never, but a near-zero effect of playing more than 3 hours. Similarly, over 3 hours of playing first-person shooter games was associated with a negative (non-significant) effect on mental rotation, but the effects at 1–30 (non-significant) and 31–120 minutes (β = 0.54, 95% CI: 0.04, 1.06) were positive. Finally, although the model (*p* = 0.077) and individual terms were not significant, the coefficients for sustained attention and overall gaming duration were positive for less than 120 minutes of play (β = 0.13-0.16) but negative (β = -0.27) for 3+ hours.

## Discussion

In this large prospective cohort of adolescents, overall videogaming duration at age 11-12 years was not longitudinally associated with subsequent cognitive development across executive functioning, visual attention, mental rotation or sustained attention. Several genre-specific longitudinal associations were observed despite the absence of associations for overall gaming duration, reinforcing the importance of considering videogaming as a heterogeneous activity rather than a single exposure. Adolescents who reported occasionally playing first-person shooter games demonstrated higher mental rotation scores than those playing never or often, and higher mental rotation scores were also observed for adolescents playing strategy games occasionally and often. Platform gaming was associated with greater sustained attention; however, FPS gaming was associated with poorer sustained attention at follow-up, and frequent role-playing game use showed a similar association.

Meta-analytic evidence has suggested that action videogames are associated with the best cognitive outcomes(1, 8). In contrast, our study observed very little association with playing action games, however this may be due to the range and precision of categories listed. For example, a widely cited study included *Halo* in their ‘action game’ category, which our participants may have categorised under ‘first-person shooter’ (19). As genre is increasingly considered a key predictor in the literature (often involving participants applying their own definition to what they play) this highlights potential issues for exposure misclassification and the potential need for standardised definitions/approaches to categorisation.

Cross-sectional associations were observed between playing many genres of game and cognition, especially executive functioning, however there was no overlap between the cross-sectional and longitudinal results, with the former potentially attributable to pre-existing differences or contextual factors rather than the impact of videogaming itself. As videogaming is a behaviourally driven exposure, this highlights the importance of longitudinal studies and the need to interpret the cross-sectional literature with caution.

The most consistent longitudinal findings were observed for sustained attention, where the association between videogaming duration and cognitive performance differed by gender. Greater baseline videogaming duration predicted poorer sustained attention at follow-up in boys, whereas it predicted better visual attention in girls. Weekend gaming, rather than weekday gaming, appeared to drive both positive and negative gender-based associations more strongly than weekday gaming, suggesting that when gaming occurs is also of importance. Although another large study of English adolescents also reported differential effects of weekend versus weekday play, they found that weekday gameplay was a stronger predictor of wellbeing, although using computers and watching films and TV was more impactful on weekends(22). These findings support the hypothesis that videogaming does not have uniform cognitive effects across adolescents and that gender may moderate associations between gaming behaviour and cognitive development.

The mechanisms underlying these gender differences remain uncertain. Boys and girls differ substantially in gaming participation, preferred genres, motivations for play, and social contexts of gaming(7, 23), all of which may contribute to different experiences and effects. Positive effects on sustained attention also emerged in girls after adjusting for action, multiplayer, racing and role-player gaming, all of which girls played substantially less often in this study. Experimental evidence has previously suggested that action videogame training can reduce gender differences in visuospatial cognition, potentially because girls have less prior exposure to these types of games and therefore greater capacity to improve these skills in this way (8). Our findings similarly suggest that the cognitive consequences of videogaming may depend on prior experience and patterns of engagement. Future work incorporating repeated measures of gaming behaviour and more detailed information on what, when, why and with whom people play would help clarify these mechanisms.

Analyses also suggested that the relationship between gaming duration and cognition may be non-linear. Although continuous measures of total gaming duration were generally not associated with cognitive development, categorical analyses indicated patterns of modest benefits for executive functioning, sustained attention and mental rotation among adolescents reporting 30 minutes to 2 hours of gaming per week, whereas those playing more than three hours per week generally showed coefficients closer to zero or in the opposite direction. Similar non-linear patterns were evident across gaming genres, where occasional play was frequently associated with more favourable cognitive outcomes than frequent play. While these trends require replication, they align with previous research suggesting that moderate recreational gaming and digital technology use can coexist with healthy development, whereas heavier gaming may increasingly displace behaviours important for cognitive development such as sleep, physical activity, and face-to-face social interaction (22, 24, 25). The present study was not designed to investigate these behavioural pathways directly, but they represent important areas for future research.

This study has several strengths. It utilised a large, diverse prospective cohort with repeated assessment of cognition during a critical period of adolescent development. The longitudinal design established a temporal sequence where exposure preceded the outcome measure, which strengthens causal inference and overcomes some limitations of cross-sectional design. Multiple cognitive domains were assessed using standardised computerised tasks administered under supervised testing conditions, reducing measurement variability. The availability of detailed information on gaming duration and genre also allowed investigation of heterogeneity in gaming experiences, an aspect often overlooked in previous epidemiological studies.

Several limitations should also be considered. Videogaming behaviour was self-reported and therefore subject to recall error and potential misclassification. Although analyses adjusted for key demographic characteristics and baseline cognition, residual confounding, as with all observational studies, remains possible. Complete-case analyses reduced sample sizes for some outcomes, particularly sustained attention, where school-level random effects could not always be estimated because of singular model fits. Consequently, findings for sustained attention, although internally consistent across several analyses, should be interpreted cautiously until replicated in larger samples. Additionally, the genres of gaming were captured at baseline to analyse follow-up cognition, however this may not adequately capture changes in gaming behaviour over time, such as if the genres of gaming and specific games played evolved from baseline to follow-up. Finally, the cohort comprised UK-based adolescents attending schools in Greater London, so findings may not generalise to populations in other contexts.

Our findings suggest that broad measures of total gaming duration may obscure important differences in gaming experience. In this study, cognitive associations were domain-specific, varying according to gaming genre, participant gender, when games were played, and whether they were played moderately or excessively. These results support growing recognition that the content and context of digital engagement is extremely important and that videogaming should not be conceptualised as a homogeneous behaviour when it comes to policy and intervention. Although detrimental effects were observed for some genres and in boys, these results alone do not imply that these genres or gaming in boys is always inherently harmful at an individual-level especially given there were benefits associated with moderate play. Yet, even smaller effects can reflect meaningful and modifiable population-level impacts. Future longitudinal studies with repeated objective measures of gaming behaviour, more detailed characterisation of game content and social context, and assessment of mediating lifestyle factors will be important for establishing whether these associations are causal and identifying which specific gaming experiences, if any, are likely to lead to cognitive benefits or harms.

## Data Availability

De-identified SCAMP data and code are available on request from.

## Acknowledgements

We are grateful for the input of the SCAMP Research Challenge student team at Bishopshalt School; Andrew Faulkner, Naomi Dearing and Leonardo Pascal-Murray. We are grateful to all schools, parents, and pupils who participated in SCAMP. We also thank all past and present SCAMP research team members for their hard work, dedication, and insights. We also thank the many casual workers who have helped with the SCAMP school assessments.

## Data availability

De-identified SCAMP data and code are available on request from.

## Authors contributions

RT conceived the analysis plan, interpreted the results, and wrote and revised the manuscript. JH analysed the data, interpreted the results and revised the manuscript. NC conceived the analysis plan, interpreted the data and revised the manuscript. SRL conceived the analysis plan and revised the manuscript. CS, RBS, MSCT, and ID guided the data analysis and revised the manuscript. MBT, MSCT and ID conceptualized the study, obtained the funding and revised the manuscript. All authors have approved the manuscript for publication.

## Abbreviations

SCAMP: Study of Cognition, Adolescents and Mobile Phones
NS-SEC: The National Statistics Socio-Economic Classification
TMT: Trail Making Test
BDS: Backwards Digit Span
SWM: Spatial Working Memory
AX-CPT: AX Continuous Performance Test
IQR: Interquartile Range
CI: Confidence Interval

