## Supplementary tables for "Adolescent videogaming, cognitive development and interactions with gender in the SCAMP cohort"

### **Appendix A.** Cross-sectional associations between baseline videogaming and cognitive abilities

| Outcome | Gaming | N | Level | Beta (95% CIs) | *p* | FDR corrected *p* |
| --- | --- | --- | --- | --- | --- | --- |
| Executive functioning* | Weekly duration of play | 4818 | Hours/week | *-0.01 (-0.03, 0.00)* | *0.08* | 0.11 |
|  | Weekly duration FPS play | 4817 | Hours/week | **-0.03 (-0.05, -0.02)** | **<0.0001 ^+^** | <0.0001 |
|  | Plays FPS games | 4817 | Never | **Reference** | **<0.0001 ^+^** | **<0.0001** |
|  |  |  | Occasionally | **-0.02 (-0.07, 0.03)** |  |  |
|  |  |  | Often | **-0.13 (-0.18, -0.08)** |  |  |
|  | Plays action games | 4817 | Never | **Reference** | **<0.0001 ^+^** | **<0.0001** |
|  |  |  | Occasionally | **-0.04 (-0.09, 0.00)** |  |  |
|  |  |  | Often | **-0.15 (-0.20, -0.10)** |  |  |
|  | Plays multiplayer games | 4817 | Never | Reference | 0.65 | 0.73 |
|  |  |  | Occasionally | 0.02 (-0.03, 0.06) |  |  |
|  |  |  | Often | 0.02 (-0.03, 0.07) |  |  |
|  | Plays sports games | 4817 | Never | Reference | 0.21 | 0.25 |
|  |  |  | Occasionally | 0.04 (-0.01, 0.08) |  |  |
|  |  |  | Often | 0.00 (-0.05, 0.05) |  |  |
|  | Plays racing games | 4817 | Never | **Reference** | **<0.0001 ^+^** | **<0.0001** |
|  |  |  | Occasionally | **0.11 (0.06, 0.15)** |  |  |
|  |  |  | Often | **-0.02 (-0.07, 0.04)** |  |  |
|  | Plays roleplayer games | 4817 | Never | Reference | 0.11 | 0.15 |
|  |  |  | Occasionally | 0.01 (-0.04, 0.06) |  |  |
|  |  |  | Often | -0.06 (-0.12, 0.00) |  |  |
|  | Plays puzzle games | 4817 | Never | **Reference** | **0.0002 ^+^** | **0.0004** |
|  |  |  | Occasionally | **0.08 (0.04, 0.12)** |  |  |
|  |  |  | Often | **0.01 (-0.04, 0.06)** |  |  |
|  | Plays strategy games | 4817 | Never | **Reference** | **<0.0001 ^+^** | **0.0003** |
|  |  |  | Occasionally | **0.09 (0.04, 0.14)** |  |  |
|  |  |  | Often | **0.09 (0.03, 0.15)** |  |  |
|  | Plays flight simulator games | 4817 | Never | **Reference** | **<0.0001 ^+^** | **<0.0001** |
|  |  |  | Occasionally | **-0.05 (-0.10, -0.01)** |  |  |
|  |  |  | Often | **-0.20 (-0.27, -0.14)** |  |  |
|  | Plays platform games | 4817 | Never | Reference | 0.14 | 0.18 |
|  |  |  | Occasionally | 0.04 (0.00, 0.08) |  |  |
|  |  |  | Often | 0.01 (-0.04, 0.06) |  |  |
|  | Plays nintendo games |  | Never | **Reference** | **0.0025 ^+^** | **0.0051** |
|  |  |  | Occasionally | **0.04 (0.00, 0.08)** |  |  |
|  |  |  | Often | **-0.05 (-0.09, 0.00)** |  |  |
|  | Plays other games |  | Never | **Reference** | **0.0003 ^+^** | **0.0004** |
|  |  |  | Occasionally | **0.07 (0.03, 0.11)** |  |  |
|  |  |  | Often | **-0.03 (-0.08, 0.03)** |  |  |
| Visual attention* | Weekly duration of play | 3534 | Hours/week | 0.01 (-0.02, 0.04) | 0.47 | 0.84 |
|  | Weekly duration FPS play | 3534 | Hours/week | *0.03 (0.00, 0.06)* | *0.067* | 0.67 |
|  | Plays FPS games | 3534 | Never | Reference | 0.15 | 0.79 |
|  |  |  | Occasionally | 0.09 (0.00, 0.19) |  |  |
|  |  |  | Often | 0.07 (-0.04, 0.19) |  |  |
|  | Plays action games | 3534 | Never | Reference | 0.88 | 0.92 |
|  |  |  | Occasionally | 0.02 (-0.07, 0.11) |  |  |
|  |  |  | Often | 0.02 (-0.09, 0.13) |  |  |
|  | Plays multiplayer games | 3534 | Never | **Reference** | **0.0011 ^+^** | **0.023** |
|  |  |  | Occasionally | **0.18 (0.08, 0.28)** |  |  |
|  |  |  | Often | **0.03 (-0.07, 0.14)** |  |  |
|  | Plays sports games | 3534 | Never | Reference | 0.26 | 0.84 |
|  |  |  | Occasionally | 0.08 (-0.02, 0.17) |  |  |
|  |  |  | Often | 0.02 (-0.09, 0.12) |  |  |
|  | Plays racing games | 3534 | Never | Reference | 0.44 | 0.84 |
|  |  |  | Occasionally | -0.06 (-0.15, 0.03) |  |  |
|  |  |  | Often | -0.03 (-0.15, 0.09) |  |  |
|  | Plays roleplayer games | 3534 | Never | Reference | 0.73 | 0.84 |
|  |  |  | Occasionally | -0.04 (-0.14, 0.07) |  |  |
|  |  |  | Often | -0.03 (-0.16, 0.09) |  |  |
|  | Plays puzzle games | 3534 | Never | Reference | 0.99 | 0.99 |
|  |  |  | Occasionally | -0.01 (-0.09, 0.08) |  |  |
|  |  |  | Often | 0.00 (-0.11, 0.11) |  |  |
|  | Plays strategy games | 3534 | Never | Reference | 0.16 | 0.79 |
|  |  |  | Occasionally | -0.03 (-0.13, 0.07) |  |  |
|  |  |  | Often | -0.12 (-0.25, 0.00) |  |  |
|  | Plays flight simulator games | 3534 | Never | Reference | 0.58 | 0.84 |
|  |  |  | Occasionally | 0.04 (-0.06, 0.14) |  |  |
|  |  |  | Often | 0.06 (-0.09, 0.21) |  |  |
|  | Plays platform games | 3534 | Never | Reference | 0.61 | 0.84 |
|  |  |  | Occasionally | 0.03 (-0.05, 0.12) |  |  |
|  |  |  | Often | -0.01 (-0.12, 0.09) |  |  |
|  | Plays nintendo games | 3534 | Never | Reference | 0.76 | 0.84 |
|  |  |  | Occasionally | 0.03 (-0.05, 0.11) |  |  |
|  |  |  | Often | 0.02 (-0.08, 0.12) |  |  |
|  | Plays other games | 3534 | Never | Reference | 0.60 | 0.84 |
|  |  |  | Occasionally | 0.03 (-0.05, 0.11) |  |  |
|  |  |  | Often | -0.02 (-0.15, 0.10) |  |  |
| Mental rotation* | Weekly duration of play | 2822 | Hours/week | -0.01 (-0.08, 0.05) | 0.68 | 0.80 |
|  | Weekly duration FPS play | 2822 | Hours/week | -0.03 (-0.10, 0.03) | 0.27 | 0.53 |
|  | Plays FPS games | 2822 | Never | *Reference* | *0.051* | 0.24 |
|  |  |  | Occasionally | *-0.12 (-0.34, 0.09)* |  |  |
|  |  |  | Often | *-0.30 (-0.55, -0.06)* |  |  |
|  | Plays action games | 2822 | Never | **Reference** | **0.004** | *0.08* |
|  |  |  | Occasionally | **0.00 (-0.21, 0.20)** |  |  |
|  |  |  | Often | **-0.38 (-0.63, -0.14)** |  |  |
|  | Plays multiplayer games | 2822 | Never | Reference | 0.29 | 0.53 |
|  |  |  | Occasionally | 0.17 (-0.05, 0.39) |  |  |
|  |  |  | Often | -0.01 (-0.25, 0.23) |  |  |
|  | Plays sports games | 2822 | Never | **Reference** | **0.018** | 0.12 |
|  |  |  | Occasionally | **-0.11 (-0.32, 0.11)** |  |  |
|  |  |  | Often | **-0.34 (-0.57, -0.10)** |  |  |
|  | Plays racing games | 2822 | Never | Reference | 0.18 | 0.51 |
|  |  |  | Occasionally | 0.18 (-0.02, 0.38) |  |  |
|  |  |  | Often | -0.01 (-0.28, 0.26) |  |  |
|  | Plays roleplayer games | 2822 | Never | Reference | 0.6 | 0.75 |
|  |  |  | Occasionally | 0.00 (-0.23, 0.24) |  |  |
|  |  |  | Often | -0.14 (-0.41, 0.14) |  |  |
|  | Plays puzzle games | 2822 | Never | Reference | 0.25 | 0.53 |
|  |  |  | Occasionally | 0.16 (-0.03, 0.35) |  |  |
|  |  |  | Often | 0.07 (-0.17, 0.31) |  |  |
|  | Plays strategy games | 2822 | Never | Reference | 0.34 | 0.53 |
|  |  |  | Occasionally | 0.12 (-0.11, 0.34) |  |  |
|  |  |  | Often | 0.17 (-0.11, 0.45) |  |  |
|  | Plays flight simulator games | 2822 | Never | *Reference* | *0.072* | 0.24 |
|  |  |  | Occasionally | *-0.13 (-0.36, 0.09)* |  |  |
|  |  |  | Often | *-0.33 (-0.64, -0.03)* |  |  |
|  | Plays platform games | 2822 | Never | Reference | 0.96 | 0.96 |
|  |  |  | Occasionally | -0.02 (-0.21, 0.16) |  |  |
|  |  |  | Often | -0.03 (-0.26, 0.20) |  |  |
|  | Plays nintendo games | 2822 | Never | **Reference** | **0.013** | 0.12 |
|  |  |  | Occasionally | **0.01 (-0.19, 0.20)** |  |  |
|  |  |  | Often | **-0.31 (-0.53, -0.09)** |  |  |
|  | Plays other games | 2822 | Never | *Reference* | *0.06* | 0.24 |
|  |  |  | Occasionally | *0.22 (0.04, 0.41)* |  |  |
|  |  |  | Often | *0.06 (-0.21, 0.33)* |  |  |
| Sustained attention | Weekly duration of play | 1448 | Hours/week | -0.02 (-0.06, 0.01) | 0.17 | 0.29 |
|  | Weekly duration FPS play | 1448 | Hours/week | -0.01 (-0.04, 0.02) | 0.59 | 0.73 |
|  | Plays FPS games | 1448 | Never | *Reference* | *0.081* | 0.20 |
|  |  |  | Occasionally | *-0.03 (-0.15, 0.07)* |  |  |
|  |  |  | Often | *-0.15 (-0.28, -0.02)* |  |  |
|  | Plays action games | 1448 | Never | **Reference** | **0.013** | 0.17 |
|  |  |  | Occasionally | **-0.10 (-0.21, 0.00)** |  |  |
|  |  |  | Often | **-0.18 (-0.32, -0.05)** |  |  |
|  | Plays multiplayer games | 1448 | Never | Reference | 0.98 | 0.98 |
|  |  |  | Occasionally | 0.01 (-0.11, 0.13) |  |  |
|  |  |  | Often | 0.01 (-0.12, 0.14) |  |  |
|  | Plays sports games | 1448 | Never | *Reference* | *0.057* | 0.20 |
|  |  |  | Occasionally | *-0.01 (-0.12, 0.10)* |  |  |
|  |  |  | Often | *-0.14 (-0.25, -0.01)* |  |  |
|  | Plays racing games | 1448 | Never | **Reference** | **0.017** | 0.17 |
|  |  |  | Occasionally | **0.14 (0.04, 0.24)** |  |  |
|  |  |  | Often | **0.10 (-0.04, 0.24)** |  |  |
|  | Plays roleplayer games | 1448 | Never | *Reference* | *0.051* | 0.20 |
|  |  |  | Occasionally | *0.13 (0.01, 0.25)* |  |  |
|  |  |  | Often | *0.13 (-0.02, 0.26)* |  |  |
|  | Plays puzzle games | 1448 | Never | Reference | 0.27 | 0.42 |
|  |  |  | Occasionally | 0.07 (-0.03, 0.17) |  |  |
|  |  |  | Often | -0.01 (-0.13, 0.12) |  |  |
|  | Plays strategy games | 1448 | Never | *Reference* | *0.077* | 0.20 |
|  |  |  | Occasionally | *0.09 (-0.03, 0.20)* |  |  |
|  |  |  | Often | *0.14 (0.00, 0.29)* |  |  |
|  | Plays flight simulator games | 1448 | Never | Reference | 0.63 | 0.74 |
|  |  |  | Occasionally | 0.05 (-0.07, 0.16) |  |  |
|  |  |  | Often | -0.03 (-0.22, 0.15) |  |  |
|  | Plays platform games | 1448 | Never | Reference | 0.17 | 0.29 |
|  |  |  | Occasionally | 0.07 (-0.02, 0.16) |  |  |
|  |  |  | Often | -0.04 (-0.16, 0.09) |  |  |
|  | Plays nintendo games | 1448 | Never | Reference | 0.8 | 0.85 |
|  |  |  | Occasionally | 0.00 (-0.09, 0.10) |  |  |
|  |  |  | Often | -0.04 (-0.16, 0.08) |  |  |
|  | Plays other games | 1448 | Never | **Reference** | **0.042** | 0.20 |
|  |  |  | Occasionally | **0.11 (0.02, 0.20)** |  |  |
|  |  |  | Often | **-0.03 (-0.19, 0.12)** |  |  |
| All models are adjusted for age + gender + ethnicity + National Statistics Socio-economic Classification (NS-SEC) + school type  *includes a random intercept for school ID  Exposure *p*-values were generated using a Chi Square test between models with and without the exposure variable.  FPS: First-person shooter  Bold: *p* < 0.05  *Italics*: *p* < 0.10  ^+^survives FDR correction | | | | | | |

### **Appendix B.** Longitudinal associations between baseline videogaming and follow-up cognition

| Outcome | Gaming | N | Level | Beta (95% CIs) | *p* | FDR corrected *p* |
| --- | --- | --- | --- | --- | --- | --- |
| Executive functioning* | Weekly duration of play at baseline | 2599 | Hours/week | 0.00 (-0.02, 0.01) | 0.57 | 0.72 |
|  | Weekly duration FPS play at baseline | 2599 | Hours/week | -0.01 (-0.02, 0.01) | 0.53 | 0.71 |
|  | Average weekly duration between timepoints | 2594 | Hours/week | -0.01 (-0.03, 0.01) | 0.34 | 0.71 |
|  | Average weekly FPS duration between timepoints | 2591 | Hours/week | -0.01 (-0.02, 0.01) | 0.53 | 0.71 |
|  | Plays FPS games | 2599 | Never | Reference | 0.16 | 0.71 |
|  |  |  | Occasionally | 0.02 (-0.03, 0.07) |  |  |
|  |  |  | Often | -0.04 (-0.10, 0.02) |  |  |
|  | Plays action games | 2599 | Never | Reference | 0.26 | 0.71 |
|  |  |  | Occasionally | 0.00 (-0.05, 0.04) |  |  |
|  |  |  | Often | -0.05 (-0.11, 0.01) |  |  |
|  | Plays multiplayer games | 2599 | Never | Reference | 0.13 | 0.71 |
|  |  |  | Occasionally | 0.01 (-0.04, 0.06) |  |  |
|  |  |  | Often | -0.05 (-0.11, 0.00) |  |  |
|  | Plays sports games | 2599 | Never | Reference | 0.72 | 0.82 |
|  |  |  | Occasionally | -0.01 (-0.06, 0.04) |  |  |
|  |  |  | Often | -0.02 (-0.08, 0.03) |  |  |
|  | Plays racing games | 2599 | Never | Reference | 0.36 | 0.71 |
|  |  |  | Occasionally | 0.02 (-0.03, 0.07) |  |  |
|  |  |  | Often | -0.03 (-0.09, 0.04) |  |  |
|  | Plays roleplayer games | 2599 | Never | Reference | 0.29 | 0.71 |
|  |  |  | Occasionally | 0.01 (-0.05, 0.06) |  |  |
|  |  |  | Often | -0.05 (-0.12, 0.02) |  |  |
|  | Plays puzzle games | 2599 | Never | Reference | 0.62 | 0.74 |
|  |  |  | Occasionally | 0.02 (-0.02, 0.07) |  |  |
|  |  |  | Often | 0.02 (-0.04, 0.08) |  |  |
|  | Plays strategy games | 2599 | Never | Reference | 0.38 | 0.71 |
|  |  |  | Occasionally | 0.03 (-0.02, 0.09) |  |  |
|  |  |  | Often | 0.02 (-0.04, 0.09) |  |  |
|  | Plays flight simulator games | 2599 | Never | Reference | 0.13 | 0.71 |
|  |  |  | Occasionally | -0.01 (-0.06, 0.04) |  |  |
|  |  |  | Often | -0.08 (-0.16, 0.00) |  |  |
|  | Plays platform games | 2599 | Never | Reference | 0.52 | 0.71 |
|  |  |  | Occasionally | 0.02 (-0.02, 0.07) |  |  |
|  |  |  | Often | 0.00 (-0.05, 0.06) |  |  |
|  | Plays nintendo games | 2599 | Never | Reference | 0.45 | 0.71 |
|  |  |  | Occasionally | 0.01 (-0.03, 0.05) |  |  |
|  |  |  | Often | -0.02 (-0.08, 0.03) |  |  |
|  | Plays other games | 2599 | Never | Reference | 0.41 | 0.71 |
|  |  |  | Occasionally | -0.01 (-0.05, 0.03) |  |  |
|  |  |  | Often | -0.04 (-0.11, 0.02) |  |  |
| Visual attention* | Weekly duration of play | 1733 | Hours/week | 0.03 (-0.02, 0.08) | 0.29 | 0.77 |
|  | Weekly duration FPS play | 1733 | Hours/week | 0.00 (-0.05, 0.05) | 0.86 | 0.97 |
|  | Average weekly duration between timepoints | 1719 | Hours/week | 0.02 (-0.04, 0.09) | 0.45 | 0.77 |
|  | Average weekly FPS duration between timepoints | 1719 | Hours/week | 0.04 (-0.01, 0.10) | 0.12 | 0.57 |
|  | Plays FPS games | 1733 | Never | Reference | 0.61 | 0.81 |
|  |  |  | Occasionally | 0.05 (-0.10, 0.21) |  |  |
|  |  |  | Often | 0.09 (-0.10, 0.27) |  |  |
|  | Plays action games | 1733 | Never | Reference | 0.11 | 0.57 |
|  |  |  | Occasionally | 0.07 (-0.07, 0.22) |  |  |
|  |  |  | Often | 0.20 (0.01, 0.38) |  |  |
|  | Plays multiplayer games | 1733 | Never | Reference | 0.97 | 0.97 |
|  |  |  | Occasionally | -0.01 (-0.17, 0.15) |  |  |
|  |  |  | Often | 0.01 (-0.16, 0.19) |  |  |
|  | Plays sports games | 1733 | Never | Reference | 0.71 | 0.87 |
|  |  |  | Occasionally | -0.05 (-0.19, 0.10) |  |  |
|  |  |  | Often | -0.07 (-0.23, 0.10) |  |  |
|  | Plays racing games | 1733 | Never | Reference | 0.27 | 0.77 |
|  |  |  | Occasionally | -0.11 (-0.25, 0.02) |  |  |
|  |  |  | Often | -0.06 (-0.26, 0.13) |  |  |
|  | Plays roleplayer games | 1733 | Never | Reference | 0.10 | 0.57 |
|  |  |  | Occasionally | 0.06 (-0.11, 0.23) |  |  |
|  |  |  | Often | 0.22 (0.02, 0.42) |  |  |
|  | Plays puzzle games | 1733 | Never | Reference | *0.056* | 0.57 |
|  |  |  | Occasionally | 0.11 (-0.02, 0.25) |  |  |
|  |  |  | Often | -0.06 (-0.23, 0.11) |  |  |
|  | Plays strategy games | 1733 | Never | Reference | 0.73 | 0.87 |
|  |  |  | Occasionally | 0.06 (-0.10, 0.21) |  |  |
|  |  |  | Often | -0.02 (-0.22, 0.19) |  |  |
|  | Plays flight simulator games | 1733 | Never | Reference | 0.35 | 0.77 |
|  |  |  | Occasionally | 0.06 (-0.10, 0.21) |  |  |
|  |  |  | Often | 0.17 (-0.07, 0.41) |  |  |
|  | Plays platform games | 1733 | Never | **Reference** | **0.049** | 0.57 |
|  |  |  | Occasionally | **0.16 (0.03, 0.28)** |  |  |
|  |  |  | Often | **0.03 (-0.13, 0.20)** |  |  |
|  | Plays nintendo games | 1733 | Never | Reference | 0.36 | 0.77 |
|  |  |  | Occasionally | 0.08 (-0.05, 0.21) |  |  |
|  |  |  | Often | -0.03 (-0.19, 0.13) |  |  |
|  | Plays other games | 1733 | Never | Reference | 0.97 | 0.97 |
|  |  |  | Occasionally | 0.01 (-0.12, 0.13) |  |  |
|  |  |  | Often | -0.02 (-0.23, 0.19) |  |  |
| Mental rotation* | Weekly duration of play | 845 | Hours/week | -0.07 (-0.22, 0.06) | 0.28 | 0.75 |
|  | Weekly duration FPS play | 845 | Hours/week | -0.06 (-0.19, 0.06) | 0.31 | 0.75 |
|  | Average weekly duration between timepoints | 838 | Hours/week | -0.03 (-0.20, 0.13) | 0.69 | 0.89 |
|  | Average weekly FPS duration between timepoints | 838 | Hours/week | 0.01 (-0.13, 0.15) | 0.90 | 0.97 |
|  | Plays FPS games | 845 | Never | **Reference** | **0.032** | 0.38 |
|  |  |  | Occasionally | **0.45 (0.04, 0.86)** |  |  |
|  |  |  | Often | **-0.13 (-0.61, 0.34)** |  |  |
|  | Plays action games | 845 | Never | Reference | 0.63 | 0.89 |
|  |  |  | Occasionally | -0.05 (-0.44, 0.33) |  |  |
|  |  |  | Often | 0.20 (-0.30, 0.68) |  |  |
|  | Plays multiplayer games | 845 | Never | Reference | 0.72 | 0.89 |
|  |  |  | Occasionally | 0.00 (-0.43, 0.45) |  |  |
|  |  |  | Often | 0.19 (-0.26, 0.62) |  |  |
|  | Plays sports games | 845 | Never | Reference | 0.38 | 0.75 |
|  |  |  | Occasionally | -0.17 (-0.57, 0.21) |  |  |
|  |  |  | Often | -0.30 (-0.76, 0.14) |  |  |
|  | Plays racing games | 845 | Never | Reference | 0.97 | 0.97 |
|  |  |  | Occasionally | 0.05 (-0.32, 0.41) |  |  |
|  |  |  | Often | 0.02 (-0.50, 0.51) |  |  |
|  | Plays roleplayer games | 845 | Never | Reference | 0.10 | 0.61 |
|  |  |  | Occasionally | 0.39 (-0.05, 0.81) |  |  |
|  |  |  | Often | 0.43 (-0.10, 0.94) |  |  |
|  | Plays puzzle games | 845 | Never | Reference | 0.74 | 0.89 |
|  |  |  | Occasionally | 0.15 (-0.22, 0.50) |  |  |
|  |  |  | Often | 0.08 (-0.37, 0.51) |  |  |
|  | Plays strategy games | 845 | Never | **Reference** | **0.019** | 0.38 |
|  |  |  | Occasionally | **0.42 (0.00, 0.82)** |  |  |
|  |  |  | Often | **0.65 (0.12, 1.17)** |  |  |
|  | Plays flight simulator games | 845 | Never | Reference | 0.17 | 0.75 |
|  |  |  | Occasionally | -0.38 (-0.80, 0.01) |  |  |
|  |  |  | Often | -0.08 (-0.70, 0.52) |  |  |
|  | Plays platform games | 845 | Never | Reference | 0.78 | 0.90 |
|  |  |  | Occasionally | -0.12 (-0.46, 0.22) |  |  |
|  |  |  | Often | -0.08 (-0.52, 0.34) |  |  |
|  | Plays nintendo games | 845 | Never | Reference | 0.34 | 0.75 |
|  |  |  | Occasionally | -0.10 (-0.45, 0.25) |  |  |
|  |  |  | Often | -0.30 (-0.71, 0.10) |  |  |
|  | Plays other games | 845 | Never | Reference | 0.60 | 0.89 |
|  |  |  | Occasionally | 0.09 (-0.24, 0.43) |  |  |
|  |  |  | Often | -0.16 (-0.68, 0.34) |  |  |
| Sustained attention | Weekly duration of play | 366 | Hours/week | -0.04 (-0.11, 0.04) | 0.37 | 0.50 |
|  | Weekly duration FPS play | 366 | Hours/week | **-0.07 (-0.13, 0.00)** | **0.048** | 0.16 |
|  | Average weekly duration between timepoints | 364 | Hours/week | *-0.08 (-0.18, 0.01)* | *0.08* | 0.21 |
|  | Average weekly FPS duration between timepoints | 364 | Hours/week | -0.04 (-0.12, 0.04) | 0.31 | 0.43 |
|  | Plays FPS games | 366 | Never | **Reference** | **0.022** | 0.11 |
|  |  |  | Occasionally | **0.00 (-0.20, 0.21)** |  |  |
|  |  |  | Often | **-0.32 (-0.56, -0.07)** |  |  |
|  | Plays action games | 366 | Never | Reference | 0.10 | 0.24 |
|  |  |  | Occasionally | -0.14 (-0.33, 0.05) |  |  |
|  |  |  | Often | -0.25 (-0.50, 0.00) |  |  |
|  | Plays multiplayer games | 366 | Never | Reference | 0.16 | 0.29 |
|  |  |  | Occasionally | 0.08 (-0.13, 0.29) |  |  |
|  |  |  | Often | -0.18 (-0.41, 0.05) |  |  |
|  | Plays sports games | 366 | Never | Reference | 0.90 | 0.92 |
|  |  |  | Occasionally | 0.00 (-0.18, 0.19) |  |  |
|  |  |  | Often | 0.05 (-0.18, 0.27) |  |  |
|  | Plays racing games | 366 | Never | Reference | 0.23 | 0.35 |
|  |  |  | Occasionally | 0.15 (-0.03, 0.33) |  |  |
|  |  |  | Often | 0.02 (-0.23, 0.28) |  |  |
|  | Plays roleplayer games | 366 | Never | **Reference** | **0.02** | 0.11 |
|  |  |  | Occasionally | **0.14 (-0.07, 0.34)** |  |  |
|  |  |  | Often | **-0.28 (-0.54, -0.02)** |  |  |
|  | Plays puzzle games | 366 | Never | Reference | 0.43 | 0.51 |
|  |  |  | Occasionally | 0.06 (-0.11, 0.23) |  |  |
|  |  |  | Often | 0.15 (-0.07, 0.37) |  |  |
|  | Plays strategy games | 366 | Never | Reference | 0.42 | 0.51 |
|  |  |  | Occasionally | 0.11 (-0.10, 0.32) |  |  |
|  |  |  | Often | 0.13 (-0.12, 0.38) |  |  |
|  | Plays flight simulator games | 366 | Never | Reference | 0.92 | 0.92 |
|  |  |  | Occasionally | 0.01 (-0.19, 0.21) |  |  |
|  |  |  | Often | 0.06 (-0.25, 0.37) |  |  |
|  | Plays platform games | 366 | Never | **Reference** | **0.012** | 0.11 |
|  |  |  | Occasionally | **0.15 (-0.01, 0.31)** |  |  |
|  |  |  | Often | **0.32 (0.10, 0.53)** |  |  |
|  | Plays nintendo games | 366 | Never | Reference | 0.15 | 0.29 |
|  |  |  | Occasionally | -0.02 (-0.19, 0.15) |  |  |
|  |  |  | Often | 0.19 (-0.03, 0.40) |  |  |
|  | Plays other games | 366 | Never | Reference | 0.78 | 0.85 |
|  |  |  | Occasionally | 0.06 (-0.10, 0.22) |  |  |
|  |  |  | Often | 0.04 (-0.25, 0.33) |  |  |
| All models are adjusted for baseline cognitive score + timepoint difference+ baseline age + baseline ethnicity +baseline gender+ baseline National Statistics Socio-economic Classification (NS-SEC) + school type  Gaming is at baseline unless otherwise stated.  Exposure *p-*values were generated using a ChiSq test between models with and without the exposure variable.  *includes a random intercept for school ID  FPS: First-person shooter  Bold: *p* < 0.05  *Italics*: *p*<0.10  ^+^survives FDR correction | | | | | | |

### **Appendix C.** Effect modification of longitudinal associations by gender

|  |  |  |  | **Males** | | **Females** | |
| --- | --- | --- | --- | --- | --- | --- | --- |
| **Outcome** | **Baseline Gaming** | **Gender interaction effect *p*** | **Adjustment for Game genre** | **Beta (95% CI)** | **p** | **Beta (95% CI)** | **p** |
| Visual attention* | Weekly duration of play | **0.037** | None | -0.01 (-0.07, 0.06)  N: 694 | 0.86 | **0.10 (0.01, 0.18)**  **N: 1039** | **0.029** |
|  |  |  | FPS | -0.03 (-0.10, 0.04)  N:694 | 0.41 | **0.10 (0.01, 0.19)**  **N: 1039** | **0.024** |
|  |  |  | Action | -0.02 (-0.09, 0.05)  N:694 | 0.54 | *0.08 (-0.01, 0.17)*  *N: 1039* | *0.087* |
|  |  |  | Multiplayer | -0.01 (-0.08, 0.06)  N:694 | 0.74 | **0.11 (0.01, 0.20)**  **N: 1039** | **0.025** |
|  |  |  | Sport | -0.01 (-0.07, 0.06)  N:694 | 0.87 | **0.10 (0.01, 0.19)**  **N: 1039** | **0.027** |
|  |  |  | Racing | -0.01 (-0.07, 0.06)  N:694 | 0.89 | **0.12 (0.03, 0.20) N: 1039** | **0.011 ^+^** |
|  |  |  | Roleplayer | -0.02 (-0.09, 0.05)  N:694 | 0.56 | 0.08 (-0.01, 0.17)  N: 1039 | 0.10 |
|  |  |  | Strategy | -0.01 (-0.07, 0.06)  N:694 | 0.84 | **0.10 (0.01, 0.19)**  **N: 1039** | **0.029** |
| Sustained attention | Weekly duration of play | **0.0051** | None | **-0.13 (-0.25, -0.01)**  **N: 158** | **0.039** | *0.10 (-0.01, 0.21)*  *N: 208* | *0.084* |
|  |  |  | FPS | -0.10 (-0.23, 0.02)  N: 158 | 0.11 | 0.09 (-0.02, 0.21)  N: 208 | 0.10 |
|  |  |  | Action | *-0.12 (-0.25, 0.00)*  *N: 158* | *0.057* | **0.13 (0.01, 0.24)**  **N: 208** | **0.031** |
|  |  |  | Multiplayer | **-0.13 (-0.26, 0.00)**  **N: 158** | **0.049** | **0.17 (0.05, 0.29)**  **N: 208** | **0.005 ^+^** |
|  |  |  | Sport | *-0.12 (-0.24, 0.01)*  *N: 158* | *0.062* | *0.10 (-0.01, 0.22)*  *N: 208* | *0.074* |
|  |  |  | Racing | **-0.13 (-0.25, -0.01)**  **N: 158** | **0.037** | **0.11 (0.00, 0.22)**  **N: 208** | **0.044** |
|  |  |  | Roleplayer | *-0.11 (-0.23, 0.02)*  *N: 158* | *0.095* | **0.13 (0.01, 0.25)**  **N: 208** | **0.030** |
|  |  |  | Strategy | **-0.13 (-0.25, -0.01)**  **N:158** | **0.03** | *0.11 (-0.01, 0.23)*  *N: 208* | *0.060* |
| All models are adjusted for baseline cognitive score + timepoint difference+ baseline age + baseline gender + baseline ethnicity + baseline National Statistics Socio-economic Classification (NS-SEC) + school type  Exposure *p*-values were generated using a ChiSq test between models with and without the exposure variable.  *includes a random intercept for school ID  FPS: First-person shooter  **Bold:** *p* < 0.05  *Italics*: *p*<0.10  ^+^survives FDR correction | | | | | | | |

### **Appendix E:** Weekend and weekday average videogaming duration at baseline (hours per week) and cognitive performance at follow-up in adolescence

| **Outcome** | **Predictor** | **N** | **β (95% CI)** | ***p*** | ***Gender interaction***  ***p*** | ***Boys*** | ***p*** | ***Girls*** | ***p*** |
| --- | --- | --- | --- | --- | --- | --- | --- | --- | --- |
| **Executive functioning** | Weekday gaming | 2599 | -0.01 (-0.02, 0.01) | 0.45 | 0.89 |  |  |  |  |
|  | Weekend gaming | 2599 | -0.01 (-0.02, 0.01) | 0.95 | 0.81 |  |  |  |  |
| **Visual attention (enumeration)** | Weekday gaming | 1733 | 0.02 (-0.03, 0.09) | 0.42 | *0.055* |  |  |  |  |
|  | Weekend gaming | 1733 | 0.02 (-0.02, 0.07) | 0.18 | **0.034** | 0.00 (-0.05, 0.05)  N = 694 | 0.91 | **0.08 (0.01, 0.14)**  **N=1039** | **0.018** |
| **Mental rotation** | Weekday gaming | 845 | -0.09 (-0.24, 0.05) | 0.20 | 0.53 |  |  |  |  |
|  | Weekend gaming | 845 | -0.02 (-0.13, 0.08) | 0.62 | 0.52 |  |  |  |  |
| **Sustained attention** | Weekday gaming | 364 | -0.02 (-0.10, 0.07) | 0.66 | **0.013** | -0.09 (-0.23, 0.04)  N = 158 | 0.16 | 0.11 (-0.02, 0.23)  N = 208 | 0.086 |
|  | Weekend gaming | 364 | -0.04 (-0.09, 0.01) | 0.15 | **0.005** | **-0.11 (-0.19, -0.03)**  **N=158** | **0.009** | 0.06 (-0.02, 0.14)  N=208 | 0.14 |
| **Bold: *p* < 0.05**  *Italics: p < 0.10*  Exposure *p*-values were generated using a ChiSq test between models with and without the exposure variable.  All models are mixed effects models, except for sustained attention, which only used fixed effects.  Models were adjusted for cognitive score at baseline, time difference between timepoints, gender, age, ethnicity, socioeconomic status, school type (fixed effects) and school (random effect). | | | | | | | | | |

### **Appendix F:** Association between categorical weekly hours of gaming at baseline and subsequent cognitive development

| **Outcome** |  | ***All gaming*** | | | ***First-person shooter gaming*** | | |
| --- | --- | --- | --- | --- | --- | --- | --- |
|  | **Categorical Predictor** | **N** | **β (95% CI)** | ***p*** | **N** | **β (95% CI)** | ***p*** |
| **Executive functioning** |  | 2599 |  | *0.068* | 2599 |  | 0.79 |
|  | Never | 183 | Reference |  | 1631 | Reference |  |
|  | 1-30 min | 1352 | 0.07 (-0.01, 0.15) |  | 419 | -0.02 (-0.08, 0.04) |  |
|  | 31 min-2 hrs | 848 | 0.09 (0.01, 0.17) |  | 357 | -0.03 (-0.09, 0.04) |  |
|  | 3 hrs or more | 216 | 0.02 (-0.08, 0.13) |  | 192 | -0.02 (-0.11, 0.06) |  |
| **Visual attention (enumeration)** |  | 1733 |  | 0.96 | 1733 |  | 0.38 |
|  | Never | 121 | Reference |  | 1096 | Reference |  |
|  | 1-30 min | 913 | -0.04 (-0.27, 0.20) |  | 294 | 0.15 (-0.02, 0.31) |  |
|  | 31 min-2 hrs | 555 | 0.00 (-0.25, 0.25) |  | 227 | 0.06 (-0.13, 0.26) |  |
|  | 3 hrs or more | 144 | -0.01 (-0.32, 0.31) |  | 116 | 0.07 (-0.19, 0.33) |  |
| **Mental rotation** |  | 845 |  | 0.34 | 845 |  | *0.064* |
|  | Never | 53 | Reference |  | 525 | Reference |  |
|  | 1-30 min | 430 | -0.06 (-0.72, 0.59) |  | 129 | 0.27 (-0.18, 0.73) |  |
|  | 31 min-2 hrs | 291 | 0.18 (-0.52, 0.88) |  | 121 | 0.54 (0.04, 1.06) |  |
|  | 3 hrs or more | 71 | -0.29 (-1.17, 0.57) |  | 70 | -0.22 (-0.85, 0.40) |  |
| **Sustained attention** |  | 366 |  | *0.077* | 366 |  | **0.0088** |
|  | Never | 27 | Reference |  | 243 | **Reference** |  |
|  | 1-30 min | 196 | 0.13 (-0.16, 0.42) |  | 48 | **0.12 (-0.11, 0.35)** |  |
|  | 31 min-2 hrs | 123 | 0.16 (-0.16, 0.48) |  | 52 | **-0.21 (-0.45, 0.04)** |  |
|  | 3 hrs or more | 20 | -0.27 (-0.72, 0.18) |  | 23 | **-0.44 (-0.77, -0.12)** |  |
| **Bold: *p* < 0.05** *Italics: p < 0.10* All models are mixed effects models, except for sustained attention, which only used fixed effects.  Exposure *p-*values were generated using a ChiSq test between models with and without the exposure variable.  Models were adjusted for cognitive score at baseline, time difference between timepoints, gender, age, ethnicity, socioeconomic status, school type (fixed effects) and school (random effect). | | | | | | | |
